# Impact and Correlation of Air Quality and Climate Variables with COVID-19 Morbidity and Mortality in Dhaka, Bangladesh

**DOI:** 10.1101/2020.09.12.20193086

**Authors:** Md Riad Sarkar Pavel, Abdus Salam, Mahbuba Yesmin, Nazmul Ahsan, Shahid Uz Zaman, Farah Jeba

## Abstract

The COVID-19 pandemic unexpectedly stopped the steady life and enhanced environmental quality. To apprehend the transmission of COVID-19 and the improvement of air quality, we have studied air quality indicators (PM_2.5_, PM_10_, AQI, and NO_2_), CO_2_ emission, and climate variables (temperature, relative humidity, rainfall, and wind velocity) in the extremely polluted and densely populated Southeast Asian megacity Dhaka, Bangladesh from March to June 2020. The Kendall and Spearman correlations were chosen to test the connotation of air quality and climate variables with COVID-19 morbidity and mortality. The average concentrations of PM_2.5_, PM_10_, and CO_2_ were 65.0 ± 37.9 and 87.1 ± 52.8 µm m_-3_, and 427 ± 11.8 ppm, respectively. The average PM_2.5_ and PM_10_ drastically reduced up to 62% during COVID-19 lockdown in Dhaka comparing with March 2020 (before lockdown). Comparing with the same period in 2019, PM_2.5_ reduced up to 33.5%. The average NO_2_ concentration was 35.0 µmol m_-2_ during the lockdown period in April, whereas 175.0 µmol m_-2_ during March (before lockdown). A significant correlation was observed between COVID-19 cases and air quality indicators. A strong correlation was obtained between climate variables and the total number of COVID-19 morbidity and mortality representing a favorable condition for spreading the virus. Our study will be very expedient for policymakers to establish a mechanism for air pollution mitigation based on scientific substantiation, and also will be an essential reference for the advance research to improve urban air quality and the transmission of the SARS-CoV-2 virus in the tropical nations.

**Graphical Abstract:** 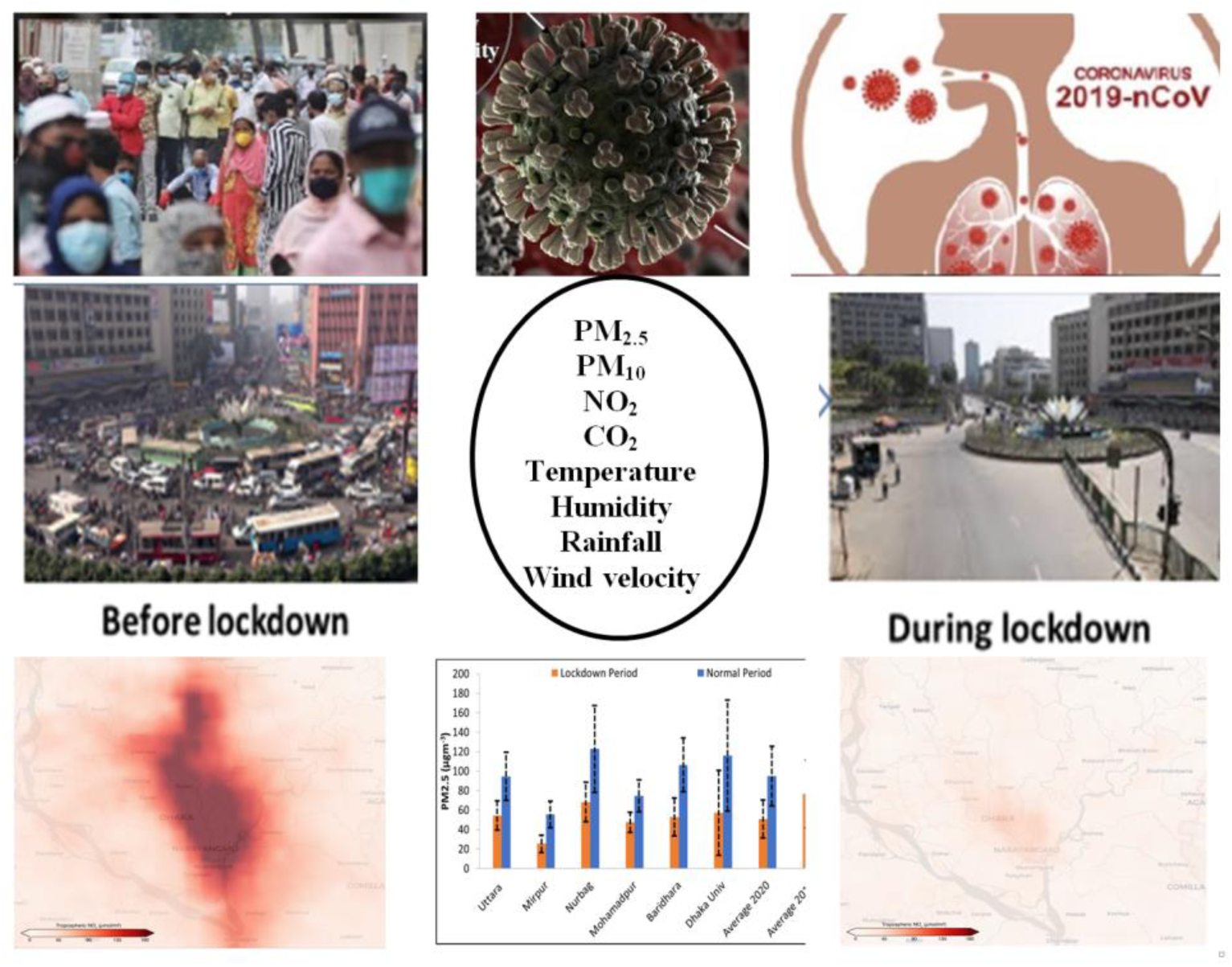

**Highlights:**

- COVID-19 lockdown drastically reduced PM_2.5_ and PM_10_ concentrations up to 62%.
- During lockdown, NO_2_ emission abridged up to 80%, CO_2_ emission dropped by 2-4%.
- Climate variables revealed strong correlation with COVID-19 morbidity and mortality.
- COVID-19 cases and mortality had significant correlation with air quality indicators.
- Knowledge from enormous improved air quality will be transmitted to the policy execution.

## Introduction

The COVID-19 pandemic has been progressing in 213 countries and territories all over the world (Hopkins, 2020). It has been a devastating attack in China, Italy, France, Spain, United States, Brazil, and many other countries (Hopkins, 2020; WHO, 2020b). In December 2019, COVID-19 started as an epidemic event in Wuhan, China having 11 million inhabitants in the central Hubei Province. People were suffering from pneumonia as the result of infection of this novel coronavirus (Li *et al*., 2020, Wu *et al*., 2020a). This virus was termed as SARS-CoV-2 on January 7, 2020, for its similarity with global epidemic Severe Acute Respiratory Syndrome (SARS) that occurred between 2002 and 2003. The severe respiratory complications are causing caused the epidemic and officially termed as COVID-19 on February 11, 2020 (Xu *et al*., 2020, WHO, 2020a). SARS-CoV-2 contains a positive-sense, single-stranded RNA and which has a size of approximately 120 nm in diameter spreading through droplets transmission (Shereen *et al*., 2020). The most severe symptoms were generally observed in older individuals having previous comorbidities, such as cardiovascular, endocrine, digestive, and respiratory diseases (Sohrabi *et al*., 2020, Wang *et al*., 2020a). Up to June 16, 2020, over 443,281 have lost their lives due to COVID-19 infection in the entire world out of 8,196,036 infections on average of 5.41 % mortality rate (https://www.worldometers.info). The virus has transmitted worldwide but, nowadays, some countries are recovering from the new transmission. However, in some regions of the world, particularly Southeast Asian countries (e.g., Bangladesh, India), the COVID-19 infection scenario is worsening day by day. Poor standard of living, limited health care facilities, high population density, worst air quality, and climate-vulnerable situation make the people of this region more endangered towards infection.

In Bangladesh, the first cases of COVID-19 were reported on March 8, 2020, and the first death of COVID-19 patient on March 18, 2020, by the Institute of Epidemiology, Disease Control and Research (IEDCR) (https://iedcr.gov.bd/). As of June 16, 2020, Bangladesh has seen a total COVID-19 infection of 94,481; total recovered of 36,262 and total death of 1,262 (mortality rate was 1.3% based on confirmed cases). The greater Dhaka region has become the epicenter of COVID-19 new cases and deaths in Bangladesh (https://iedcr.gov.bd/). The lockdown was enforced on 26th March 2020, which ended on 30th May 2020. This long-term lockdown affected industrial production, educational institutes, construction activities, small and large scale businesses. It created a huge negative impact on the economy and a large group of people is facing a crisis for their daily livings. Due to the uncontrolled situation of COVID-19, the Government of Bangladesh again instigated regional lockdown depending on the number of infected people on June 16, 2020, marked as a red, yellow, and green zone. During the lockdown, nature has got an opportunity for healing as dwellers are maintaining social distancing, quarantine at home, minimum outside activities. Therefore, we saw the blue sky in Dhaka city during the lockdown period (more than 100 days). Air quality has also substantially improved compared to the previous years in Dhaka like other cities in the World, e.g., Delhi, (−60%), Seoul (−54%), Wuhan (−44%), and Los Angeles (−31%) (IQ Air, 2020).

However, air quality was not improved significantly in the entire world. It has improved a lot in some cities but other cities it did not change at all, and the cause was not the COVID-19 lockdown only (Schiermeier, 2020). For example, a significant reduction in Los Angles, USA was not clear, either for COVID-19 precautions or falling rain during this period (Schiermeier, 2020). In the absence of traffic vehicles and control industrial activities, several air pollutants in China declined up to 90% during COVID-19 pandemic (Le *et al*., 2020) (a reduction of 22.8 μg m_-3_ NO_2_ and 18.9 μg m_-3_ PM_2.5_ were also observed in China - Chen *et al*., 2020b). Although it has not untarnished the role of aerosol particles in the transmission of the SAR-CoV-2 virus, long-term high-level exposure to air pollutants may have protracted vulnerability and mortality rates of COVID-19, (Liu *et al*., 2020).

Some studies reported the relationship between the number of confirmed COVID-19 positive cases and climate variables, e.g., temperatures, humidity (Bukhari and Jameel, 2020; Ma *et al*., 2020). Bukhari and cols, 2020 reported that the low number of positive COVID-19 confirmed cases observed in tropical countries might be explained due to their usual warm and humid environmental conditions. The regions with high relative humidity above 10 g m_-3_ could see a slowdown of COVID-19 transmissions in a short time (Bukhari and Jameel, 2020). Ma and cols, 2020 also revealed that the temperature and humidity were affecting factors related to the number of COVID-19 mortality in Wuhan, China (Ma *et al*., 2020).

Therefore, we would like to quantify the effect of lockdown on air quality parameters (PM_10_, PM_2.5_, NO_2_, and CO_2_) and also would like to elucidate the correlation between climate variables (temperature, relative humidity, rainfall, and wind velocity) and air quality indicators with COVID-19 morbidity and mortality in Dhaka, Bangladesh.

## Research Methodology

Dhaka is the capital city of Bangladesh and the center of the Government, trade, and culture of the country. Geographically, Dhaka is located in the center of Bangladesh between 23°22’30’’ N to 24°22’20” N and 89°41’6’’ E to 90°59’23” E, on the eastern banks of the Buriganga River (Fig. 1).

**Fig. 1.**
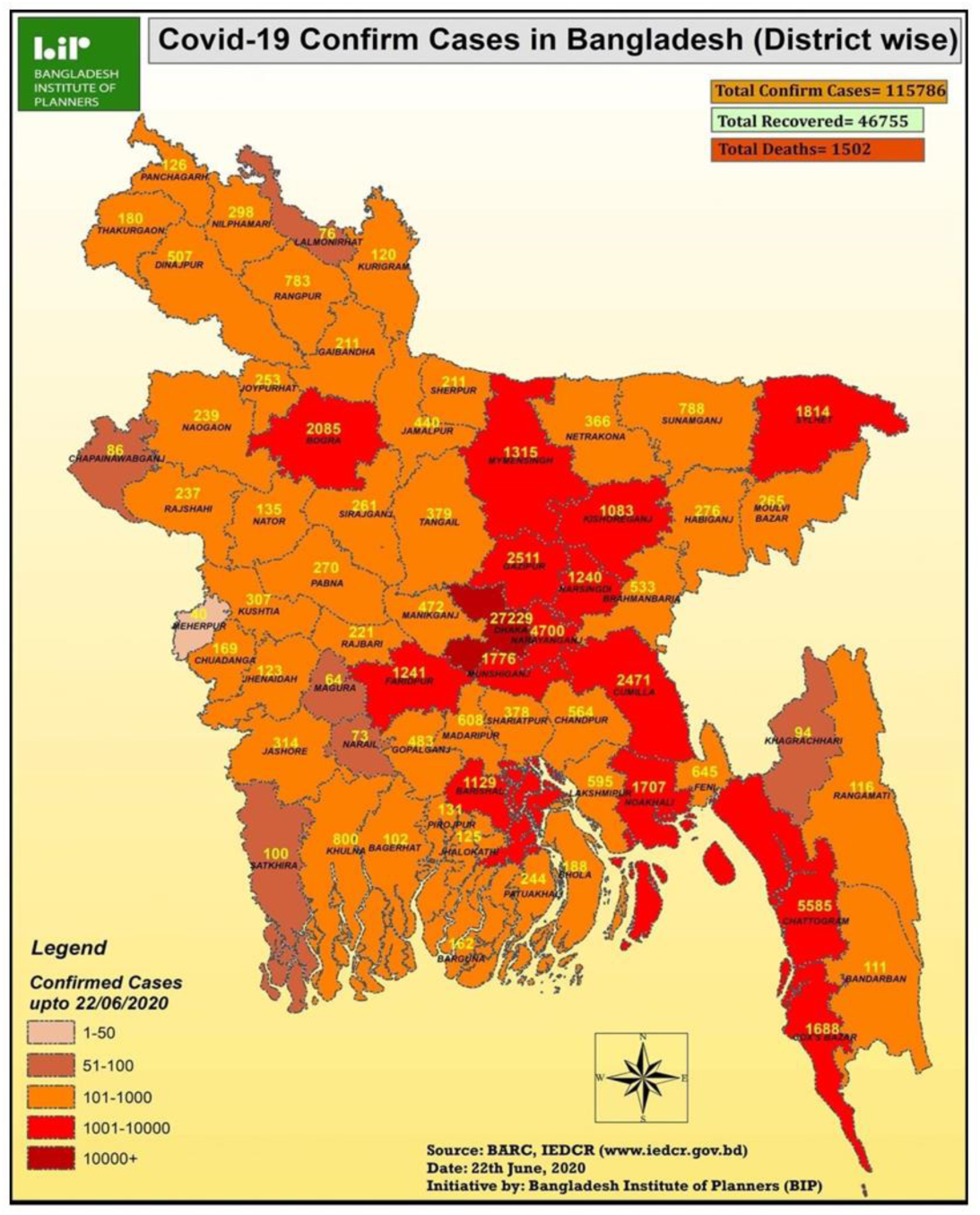
Distribution of COVID-19 confirmed cases in 64 districts of Bangladesh until June 22, 2020 (Source: Bangladesh Institute of Planners).

The climate of Bangladesh is generally hot and humid. Meteorologically, Bangladesh can be sub-divided into four seasons. Pre-monsoon (March-May), Monsoon (June-August), Post-monsoon (September-November), and winter (December-January) (Salam *et al*., 2012; Salam *et al*., 2003). The maximum summer temperature ranges from 30º C to 40º C. April is the warmest month and, January is the coldest month in general. The temperature drops to 10º C on average across the country during winter. The effect of monsoon climate is heavy rainfall. The maximum precipitation is 103.6 inches, the minimum is 46.1 inches while the average is 63.4 inches. Dhaka is the most densely populated megacity in the World with 21 million people residing within 1462.6 square kilometers. The wind direction follows the general trend of south and south-west in the pre-monsoon and north and north-west in the winter (Salam *et al*., 2003).

This study includes two stages of analysis: 1) to assess the effect of lockdown on air quality indicators (PM_2.5_ and PM_10_), CO_2_ emission, and 2) to study the correlation of climate variables (average temperature, relative humidity, rainfall, and wind velocity) and air quality parameters with COVID-19 morbidity and mortality. The Kendall and Spearman rank correlation tests were utilized to examine the correlation between variables and air quality parameters. Air quality data of Dhaka was obtained from our measurements at the Mukarram Hussain Khundker Building, University of Dhaka. Data set for COVID-19 was taken from the COVID-19 archive from the Directorate General of Health Services (DGHS) (www.dghs.gov.bd). Data on climate variables (average, relative humidity, wind speed, and rainfall) was taken from weatherforyou.com and darksky.net.

## Results and Discussion

### Overview of the Air Quality and Climate variables

The summary of PM_2.5_, PM_10_, CO_2_, temperature, relative humidity, wind velocity, and rainfall between March 8 (1st day of confirmed COVID-9 cases in Bangladesh) and June 16, 2020, in Dhaka megacity has been given in Table 1. The total average PM_2.5_ and PM_10_ were 65.0 ± 37.9 and 87.1 ± 52.8 µg m_-3_, which were about 2.6 and 1.7 times higher than WHO guideline values for 24 h average, respectively. The average PM_2.5_ was 50.8±19.6 µg m_-3_ during the lockdown and 94.9 ± 30.6 µg m_-3_ before lockdown in Dhaka megacity. The lowest average relative humidity was 39.1 %, the highest was 99.8 %, and the average rainfall was 0.23 ± 0.31 inch hour_-1_. The average CO_2_ emission was 427 ± 11.8 ppm, which is slightly lower (456 ± 63.4 ppm) than the value in January 2020.

**Table 1.** Summary of statistical values of air quality and climate variables from March 8 to June 16, 2020 in Dhaka University Area, Dhaka, Bangladesh.

| | **PM<sub>2.5</sub>**<br>($\mu\text{g m}^{-3}$) | **PM<sub>10</sub>**<br>($\mu\text{g m}^{-3}$) | **CO<sub>2</sub>**<br>(ppm) | **Temp**<br>(°C) | **Humidity**<br>(%) | **Rainfall**<br>(inch hour<sup>-1</sup>) | **Wind velocity**<br>(m h<sup>-1</sup>) |
| --- | --- | --- | --- | --- | --- | --- | --- |
| **Average** | 65.0 | 87.1 | 427.5 | 29.3 | 73.8 | 0.23 | 6.5 |
| **Median** | 57.8 | 76.8 | 425.1 | 29.4 | 76.0 | 0.08 | 6.0 |
| **S.D.** | 37.9 | 52.8 | 11.8 | 2.1 | 12.9 | 0.31 | 3.1 |
| **Max** | 207.4 | 272.6 | 465.8 | 33.6 | 99.8 | 1.59 | 18.3 |
| **Min** | 17.3 | 19.7 | 408.3 | 23.9 | 39.1 | 0.00 | 2.0 |

As control measures during COVID-19 pandemic, precautions, such as lockdown were enforced by restricting vehicle movements, closing government offices, schools, colleges, universities, shopping malls, and “stay in home order” on March 26, 2020, have been taken by the Government of Bangladesh. Due to the restrictions, the concentrations of PM_2.5_ and PM_10_ have reduced drastically up to 62% during April. The times series of daily average concentrations of PM_2.5_, PM_10_, and CO_2_ have given in Figs. 2(a), 2(b) and 2(c) mentioning the events, e.g., lockdown begins, Garments industry open, Eid festival, lockdown ends and offices, etc. The daily average global CO_2_ were also drastically reduced up to 17% with a maximum of 26% and a minimum of 4% in the early April compared to the previous year in 2019 due to the restricted activates during COVID-19 pandemic (Le Quéré *et al*., 2020). In general, the values of the air quality parameters were getting lower with the progress of the lockdown (Fig. 2). When the lockdown was curbed on May 30, 2020, we can see, the concentrations of the air quality parameters were rising slowly in Dhaka.

**Fig. 2.**
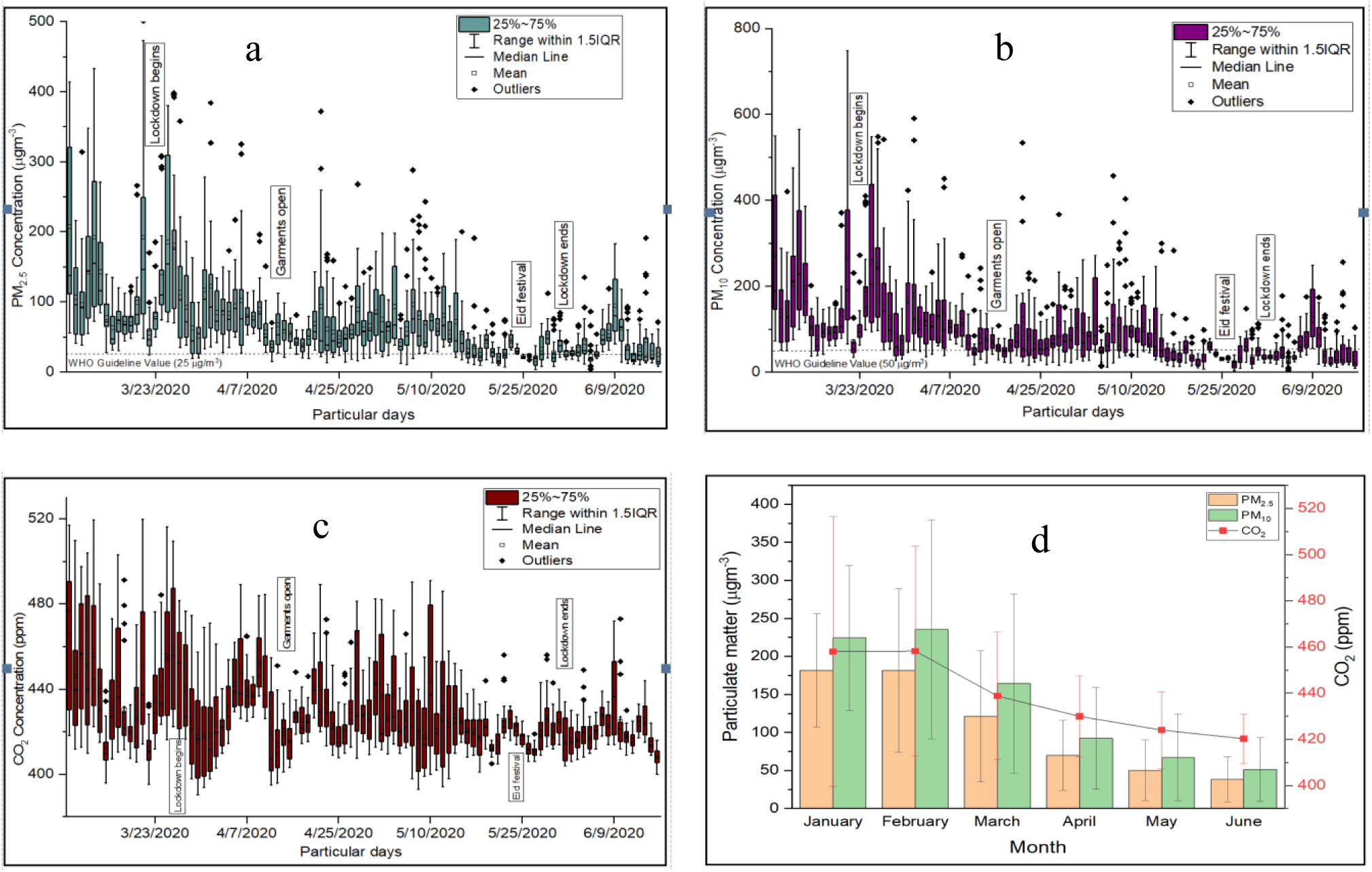
(a, b, c). The daily average concentrations of PM_2.5_, PM_10_, and CO_2_; and 2(d). Monthly average concentrations of PM_2.5_, PM_10_, and CO_2_ at Mukarram Hussain Khundker Building, University of Dhaka, Bangladesh.

The monthly average concentrations of PM_10_, PM_2.5_, and CO_2_ emission were getting lower from January through June 2020. January and February were almost similar but much higher than April, May, and June. The reduction was not only for COVID-19 precautions activities, but also rainfall from April to June (Fig. 2(d)), especially for PM_2.5_, PM_10_, and CO_2_), but NO_2_ reduction in April 2020 is only for traffic emission (Fig. 5). However, we have identified and quantified the contribution of reduction in air quality indicators due to COVID-19 lockdown by comparing it with the same period of the previous year 2019 (Fig. 3 and Fig. 4). As a case study, we have differentiated between “before lockdown” and “during lockdown” PM_2.5_ emission in Fig. 3.

**Fig. 3.**
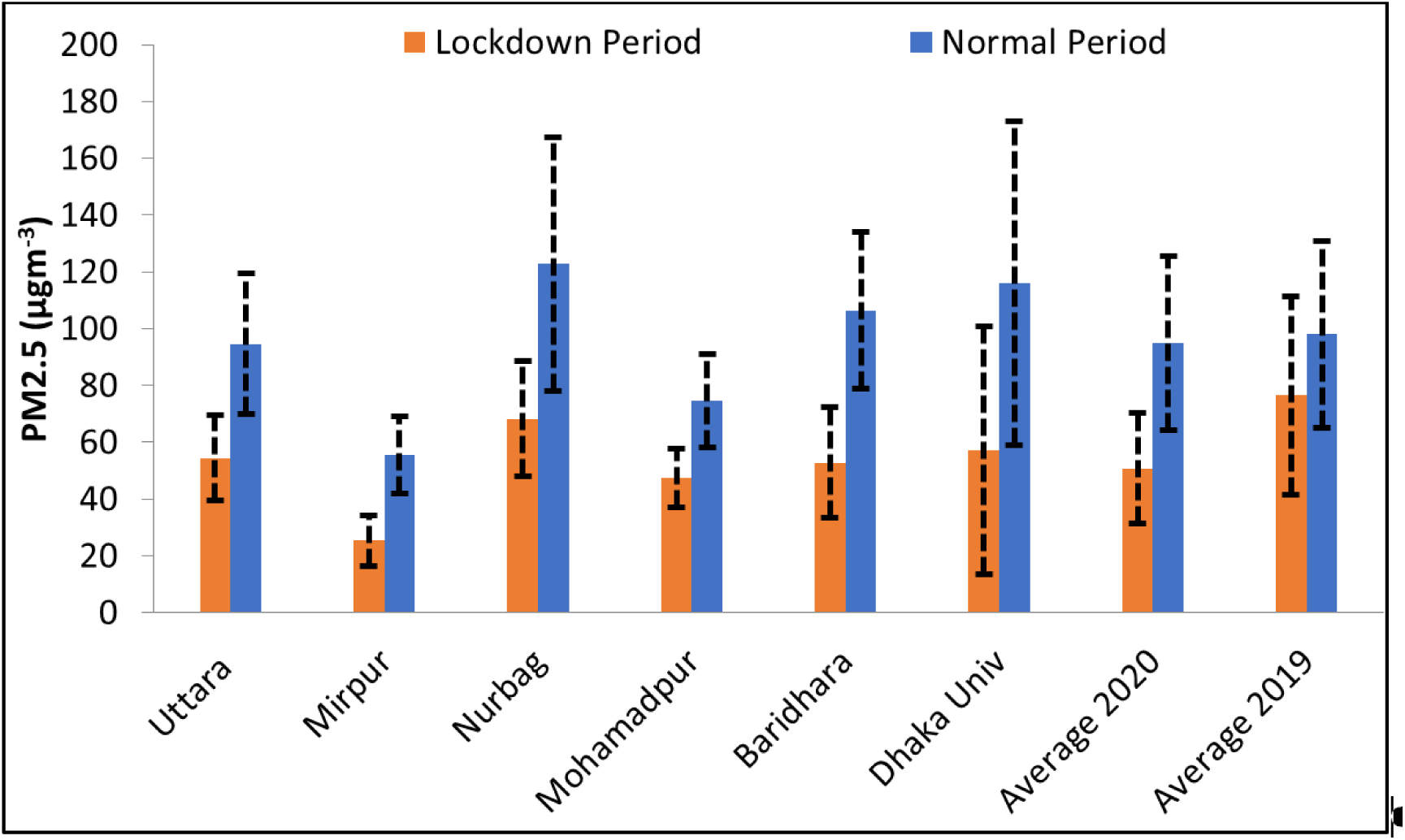
PM_2.5_ concentrations between normal (March 16-27, 2020) and lockdown (March 28-May 30, 2020) periods at five different locations in Dhaka, Bangladesh. The average PM_2.5_ concentrations of the same time period in 2019 have also been given for comparison.

**Fig. 4.**
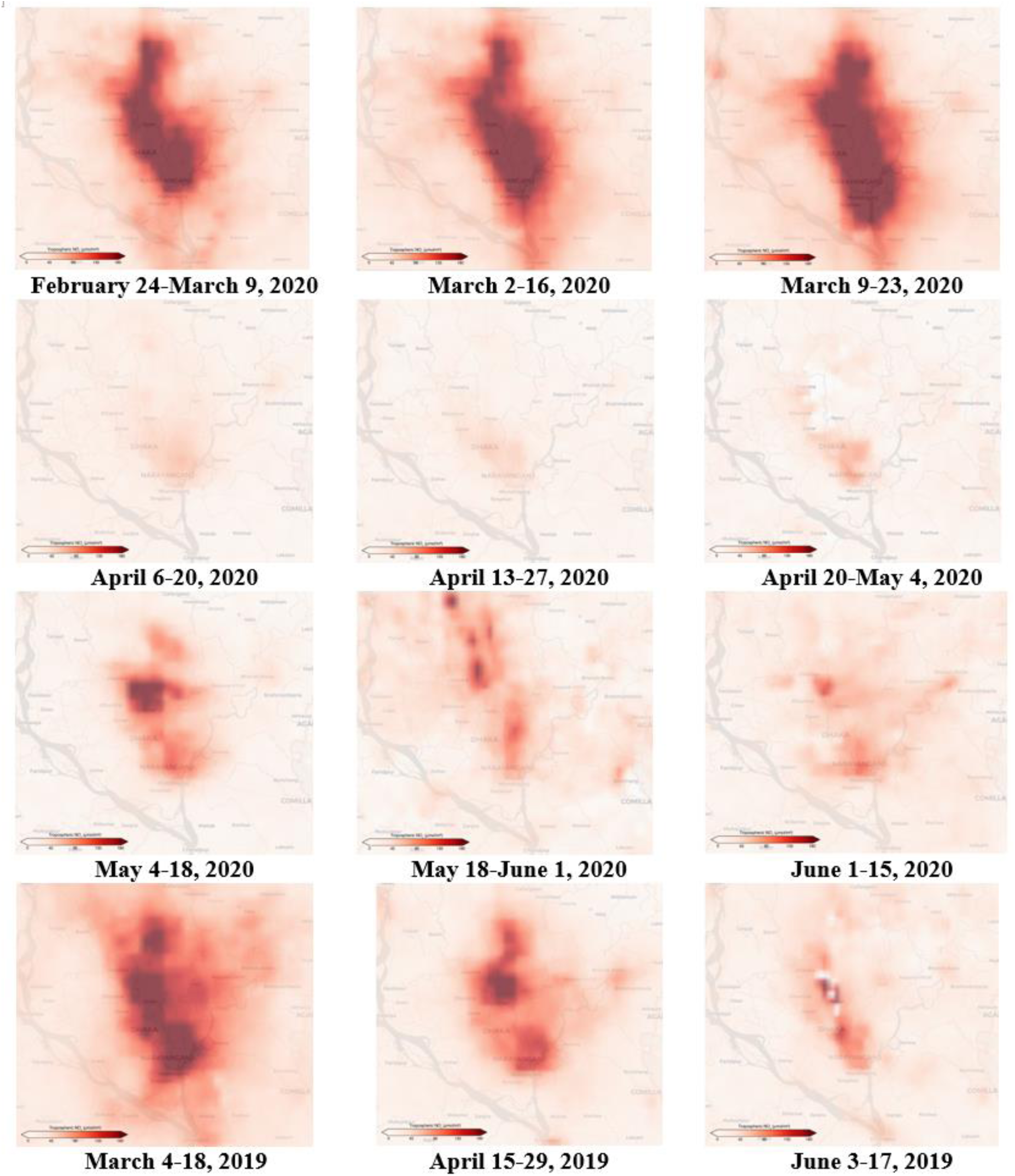
NO_2_ concentrations before COVID-19 lockdown (March 2020), during lockdown (April 2020) and after limited scale lockdown (May and June 2020), and bottom panel was from similar time period but in 2019 over Dhaka, Bangladesh (Source: European Space Agency).

Several recent papers reported that a decline in PM_2.5_ is associated with lockdown and affected COVID-19 transmission (Chauhan and Singh, 2020; Sharma *et al*., 2020). The data of the PM_2.5_ concentrations were analyzed at five different locations in Dhaka city before lockdown (March 16-27, 2020) and after lockdown (March 28-May 30, 2020) to see the variations in Dhaka city, and also made a comparison with the same period in 2019 (Fig. 3). The average PM_2.5_ decline during the lockdown period was 42.3% in Uttara, 31.6% in Mirpur, 57.4% at Nurbag, Kamrangirchar, 28.8% in Mohammadpur, 56.5% in Baridhara, 62% in Dhaka University area (Fig. 3). The overall average PM_2.5_ decreased by about 46.6% during the lockdown period. The average PM_2.5_ value during the lockdown period in 2020 also decreased by 33.5% compared to the last year (2019), but regular period values of PM_2.5_ were almost similar between 2020 and 2019 (Fig. 3).

The satellite images of NO_2_ distribution over the Dhaka city have been given in Fig. 4 at different periods before and during the lockdown, and also during limited scale lockdown in 2020; and also, for a similar period in 2019. It has clearly shown the significant reduction of NO_2_ concentration during the lockdown period in April 2020 (about 35 µmol m_-2_), whereas it was about 170 µmol m_-2_ before lockdown over Dhaka.

The comparison between April 13-27 in 2020 and April 15-29 in 2019 was also indicating a very drastic reduction of NO_2_ concentration over Dhaka. During April 2020, the vehicles were very limited in the street of Dhaka due to the complete shutdown of offices, schools, colleges, universities, constructions, industries, and also “stay in-home” order. These are the major causes of the drastic reduction of NO_2_ over Dhaka city because the foremost source of NO_2_ emissions is from traffic vehicles. The NO_2_ reduction was 40% (on average) in many cities of China, 20 - 38% in Western Europe and the USA during COVID-19 lockdown compared to the same period in 2019 (Bauwens *et al*., 2020).

## Variation of COVID-19 Morbidity and Mortality

### With Air Quality Indicators

Temporal variation of total confirmed COVID-19 cases, new cases, and daily deaths with PM_10_, PM_2.5_, and AQI in Dhaka has given in Fig. 5. The relationship between aerosol particle number and mass with SARS-CoV-2 virus has been studying but not fully understood yet. The high concentration of viral RNA has been observed when the submicron and/or super submicron particles having peaked in the aerosol size distribution curve (Liu *et al*., 2020). The relationship between COVID-19 morbidity and mortality cases and air quality index (PM_10_, PM_2.5_, and AQI) was not linear (Fig. 5). The reason may be the particle sizes - PM_2.5_ and PM_10_ are much larger compared to the size of the SARS-CoV-2 virus. However, long term exposure to a high concentration of particulate air pollution might be attributed to bronchial asthma, chronic obstructive pulmonary diseases (COPD), chronic heart diseases, which might be one of the causes of the relatively high COVID-19 death rate in Dhaka.

**Fig. 5.**
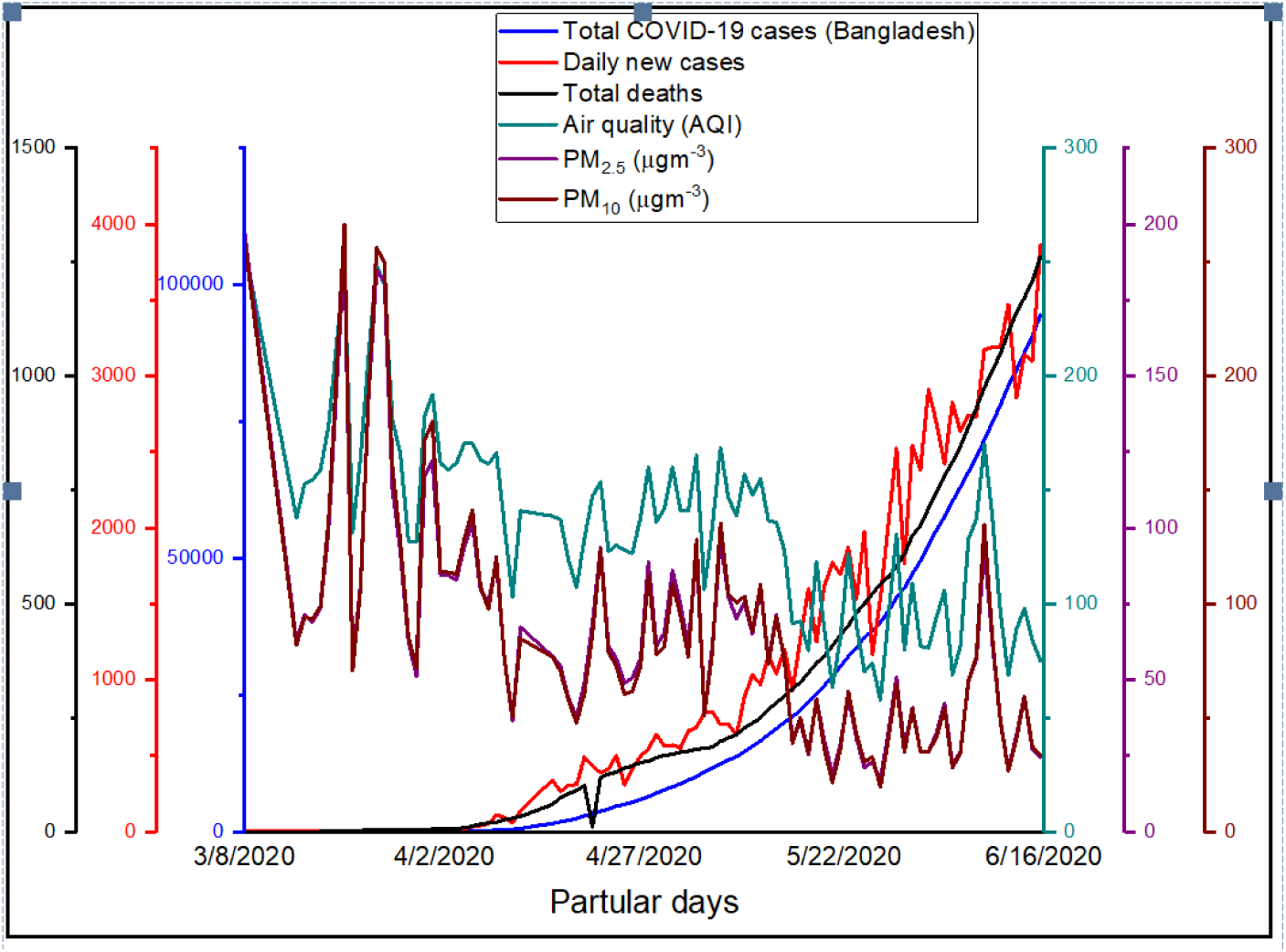
Depiction of COVID-19 cases and air quality (PM_2.5_, PM_10_, AQI) from March 8 to June 16, 2020 in Bangladesh (Source: COVID-19 related data from Directorate General of Health Services, DGHS, Bangladesh).

### With Climate variables

The variation of total COVID-19 confirmed cases, daily new cases, and total deaths with climate variables (temperate and relative humidity) in Dhaka from March 8, 2020 to June 16, 2020 has given in Fig. 6. The total number of COVID-19 confirmed cases and total deaths were increasing sharply in Dhaka. The temperature and relative humidity curve were also going up. The number of confirmed cases was 94,481 on June 16, 2020 with the total deaths of 1262. However, the relationship between climate variables and COVID-19 cases is not well understood yet, may be many factors are responsible. But a strong linear relationship has been observed for the COVID-19 total and deaths cases with temperature and relative humidity in Dhaka city (Fig. 6). Climate variables (e.g., temperature, relative humidity, wind velocity and rainfall) have a significant impact on the formation, dispersion and deposition of pollutants in the atmosphere. Ambient air pollutants with climate variables aggravated the pulmonary diseases and injured the respiratory pathways, and hence facilitated the virus to infect the human system and cause unadorned damages (Zoran *et al*., 2020).

**Fig. 6.**
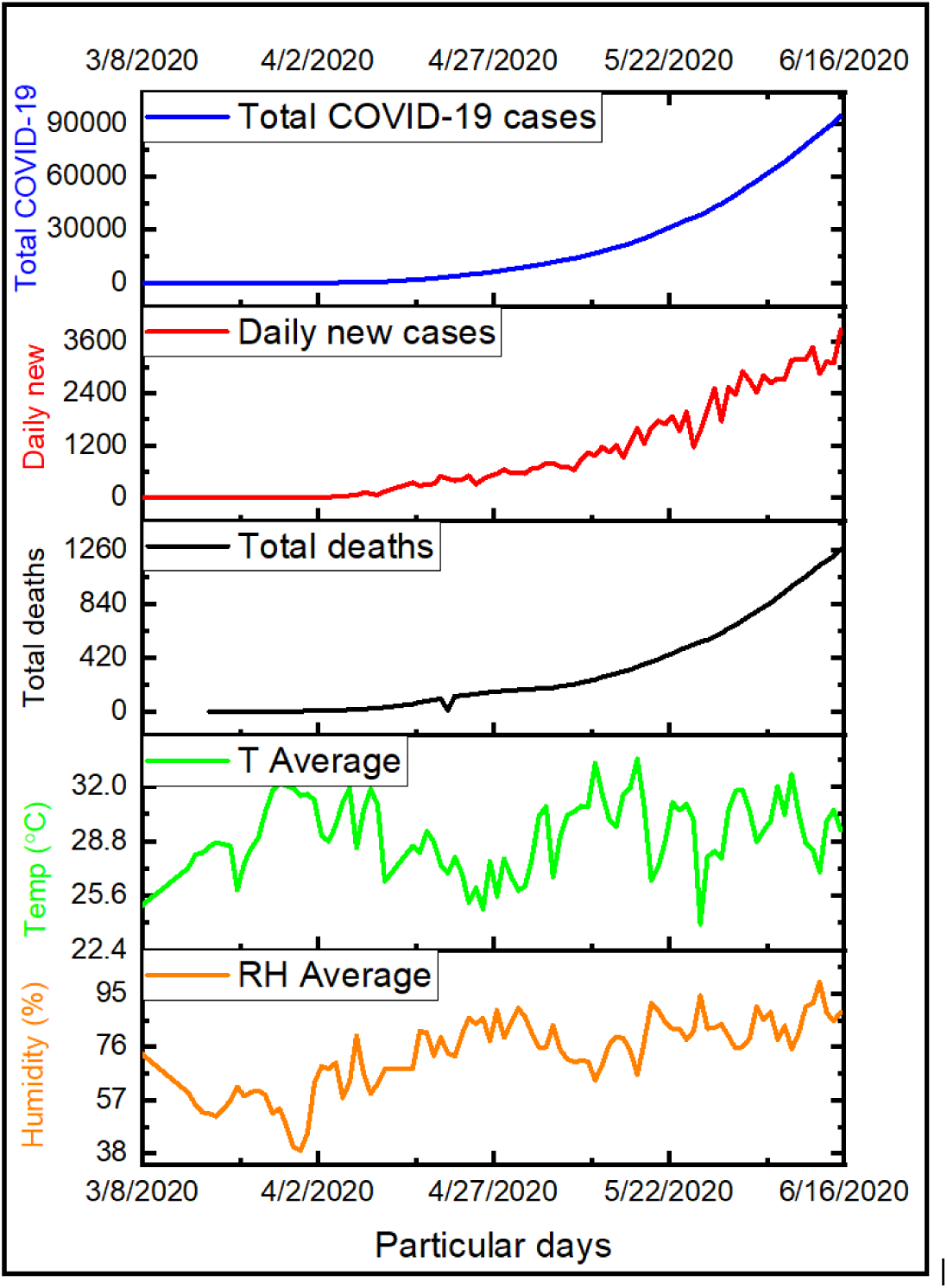
Temporal variation of total number of COVID-19 cases, daily new cases, total deaths, daily mean climate variables (average air temperature and relative humidity) in Bangladesh (Source: COVID-19 related data from Directorate General of Health Services, DGHS, Bangladesh).

### Kendall and Spearman Correlation Tests

The Kendall and Spearman correlation tests were performed between the empirical data of PM_2.5_, PM_10_, CO_2_, and AQI and climate variables (temperature, relative humidity, wind velocity, and rainfall) with COVID-19 morbidity and mortality. In both Kendall and Spearman correlation tests, we found significant negative correlation with air quality indicators (PM_2.5_, PM_10_, CO_2_, and AQI) with total confirmed COVID-19 cases, daily new cases, and mortality in Dhaka, Bangladesh (Table 2). The magnitude of the correlation coefficients of PM_2.5_, PM_10_, CO_2_, and AQI were lower in Kendall tests than the Spearman correlation. The air quality was good during lockdown period but the mortality and morbidity was high in Dhaka. Because the COVID-19 transmitted patients may be experiencing worst air quality from the previous months (January, February and March) of the year that have significant impact on their health. Patients organs (e.g., lung, heart, kidney, etc.) may already compromised by the long term exposure of the poor air quality in Dhaka. Some other factors (immune system, cofactors) may also be contributed to this significant negative correlation in Dhaka. In California, USA, Bashir et al. (2020b) also found similar results - PM_2.5_, PM_10_, SO_2_, CO, and NO_2_ have significant negative correlation with total COVID-19 morbidity and mortality.

Whereas, the higher concentrations of PM_10_ and PM_2.5_ reported a positive correlation with death rate of COVID-19 in China (Yao *et al*., 2020). Fattorini and Regili, 2020 revealed that the primary cause of COVID-19 in Northern Italy was the continuous exposure of high levels of PM_10_ and PM_2.5_ (Fattorini and Regili, 2020). As a first-order approximation, the COVID-19 related morbidity and mortality in Dhaka are significantly correlated with the high exposure of air quality parameters, especially PM_10_ and PM_2.5_ like other cities in the World (e.g., California, Wuhan, and Northern Italy). The climate variables (average temperature, relative humidity, wind velocity, and rainfall) have strong positive correlation with the total COVID-19 confirmed and daily new cases, and mortality in Dhaka.

Previous studies of Vandini *et al*., (2013) and Bashir *et al*., (2020a) reported similar results and support our findings. Shi *et al*., 2020 reported that climate indicator (temperature) serves as a driver for the COVID-19. On the other hand, Méndez-Arriaga, (2020) reported a negative association of the air temperature with the total confirmed COVID-19 cases. COVID-19 outbreak at Wuhan, China showed a strong association between diseases spread and weather conditions, with a prediction of “warm weather will play an important role in suppressing the virus.”

**Table 2.** Empirical results of correlation between air quality indicators and climate variables with total confirmed cases, new cases and mortality in Dhaka, Bangladesh.

|  | Parameters | Total Cases | New Cases | Mortality |
| --- | --- | --- | --- | --- |
| **Kendall Correlation Coefficient** | PM<sub>2.5</sub> | -0.350* | -0.234* | -0.306* |
|  | PM<sub>10</sub> | -0.329* | -0.219** | -0.265* |
|  | CO<sub>2</sub> | -0.312* | -0.315** | -0.256* |
|  | AQI | -0.362* | -0.294* | -0.278* |
|  | Average temperature | 0.161 | 0.108 | 0.202** |
|  | Relative humidity | 0.282* | 0.232* | 0.141 |
|  | Wind velocity | 0.031 | 0.060 | 0.035 |
|  | Rainfall | 0.191** | 0.159 | 0.095 |
| **Spearman Correlation Coefficient** | PM<sub>2.5</sub> | -0.523* | -0.351* | -0.438* |
|  | PM<sub>10</sub> | -0.495* | -0.418* | -0.414* |
|  | CO<sub>2</sub> | -0.426* | -0.445** | -0.376* |
|  | AQI | -0.532* | -0.430* | -0.421* |
|  | Average temperature | 0.249** | 0.158 | 0.309** |
|  | Relative humidity | 0.408* | 0.324** | 0.207 |
|  | Wind velocity | 0.063 | 0.099 | 0.053 |
|  | Rainfall | 0.303** | 0.208 | 0.205 |

\*\*\*, \*\*, \* indicate significance at levels of 10%, 5% and 1%.

However, temperature and relative humidity may play a significant role in COVID-19 transmission and also have a substantial role in the mortality rate (Auler *et al*., 2020; Poole, 2020; Sajadi *et al*., 2020; Chen *et al*., 2020a; Ma *et al*., 2020; Wang *et al*., 2020b). Other meteorological indicators such as wind velocity and rainfall also showed a significant positive correlation with COVID-19 cases in Dhaka, Bangladesh.

Dhaka, as an administrative, cultural, and economic center of Bangladesh, experiences a high influx of population from all over the country for seeking employment, education, business, and medical treatments. Moreover, many people are living not only in the slums but also in the inappropriate substandard houses with inadequate facilities and very limited medical access with the worst air quality. All these made the COVID-19 situation in Dhaka uncontrollable and hence withstand with a large number of COVID-19 confirmed and death cases for a comparatively long period. Therefore, our study will supplement new knowledge for decision-makers that air quality and climate variables are two important constraints that need to be addressed for controlling the transmission of the SARS-CoV-2 virus in the developing countries.

## Conclusion

Air quality parameters (PM_2.5_, PM_10_, AQI, NO_2_, and CO_2_) and climate variables (average temperature, relative humidity, rainfall, and wind velocity) were studied before and during the COVID-19 pandemic from March 8, 2020, to June 16, 2020, in Dhaka, Bangladesh. The study revealed the impact of COVID-19 precautions on Dhaka air quality. The Kendall and Spearman correlation tests were performed to see the correlation between COVID-19 morbidity and mortality cases with climate variables and air quality indicators. The air quality has drastically reduced in Dhaka due to COVID-19 lockdown in April 2020 (on average, about 2.5 times lower PM_2.5_ and 6 times lower NO_2_ observed than a normal period). PM_2.5_, PM_10_, CO_2_, air quality index (AQI) has no positive but a significant correlation (within 1.0%) with the number of COVID-19 confirmed cases and mortality. Average temperature, minimum temperature, and wind velocity have a strong positive correlation with COVID-19 confirmed morbidity and mortality cases in Dhaka. These findings will be helpful for policymakers to understand the mitigation mechanism of the air quality and also will be useful for future research to control the SARS-CoV-2 virus as well as air quality.

## Data Availability

All data are available upon request

## Acknowledgement

Authors acknowledge the data support to Air visual (www.iqair.com), European space agency (NO_2_ satellite images), DGHS, Bangladesh (COVID-19 related data at www.dghs.gov.bd) and for climate variables data (www.darksky.net and www.weatherforyou.com).

